# Economic Evaluation: Costing participatory learning and action cycles with women’s groups to improve feeding, care and dental hygiene for South Asian infants in London

**DOI:** 10.1101/2024.03.09.24304022

**Authors:** Yeqing Zhang, Priyanka Patil, Monica Lakhanpaul, Michelle Heys, Subarna Chakraborty, Joanna Dwardzweska, Clare H. Llewellyn, Kelley Webb-Martin, Carol Irish, Mfon Archibong, Jenny Gilmour, Phoebe Kalungi, Prof Jolene Skordis, Logan Manikam, Neha Batura, the NEON Steering Team

## Abstract

**Background:** The Nurture Early for Optimal Nutrition (NEON) programme was designed to promote equitable early childhood development by educating mothers of South Asian origin in east London on optimal feeding, care, and dental hygiene practices. This study conducts a cost analysis of the NEON programme and evaluates its financial sustainability.

**Methods:** We conducted an economic costing from the provider perspective and followed a stepdown procedure to identify all costs incurred from December 2019, the initiation of the trial, to May 2023, the completion of final evaluation and dissemination. Costs associated with start-up, implementation, and monitoring and evaluation activities are differentiated. Affordability analysis was conducted with respect to the budget of the local authorities.

**Results:** The total cost of NEON design and delivery in Newham and Towe Hamlets was £75,992 ($INT 114,445), with 45% for staff salaries, 50% for material, and 5% for capital investment. The start-up stage cost 57% while the implementation stage cost 43%. The average cost per mother participating in the programme was £409($INT 615). The total cost of trial delivery in Newham accounted for around 0.053% of the borough’s annual child development expenditure, while the total trial cost in Tower Hamlets was equivalent to 0.003% of its’ spending on children’s development.

**Conclusion:** The delivery of NEON is largely within local authorities’ budget for childhood development. The unit cost is expected to decrease when sharing costs are spread across more participants and implementing systems are validated and well developed.

## Introduction

Annually, an estimated 240 million children under the age of five worldwide are subjected to significant biological and psychosocial hazards, compromising their developmental potential ^[1]^. These hazards encompass malnutrition, exposure to violence and heavy metals, and inadequate cognitive and social-emotional stimulation ^[2]^. The initial five years of life are crucial for brain development and the formation of caregiver-child attachments, rendering this period particularly sensitive to early experiences ^[3]^. Adversity during this time can have long-term detrimental impacts on physical and psychosocial health, as well as educational and economic achievements in adulthood ^[3–7]^. In financial terms, the average annual income loss for adversely affected children is estimated at 26% ^[8]^. Furthermore, there are intergenerational consequences as the developmental deficit and income loss perpetuate a cycle of poverty ^[8]^.

Conversely, interventions during this early period have been demonstrated to yield the greatest benefits for health and development ^[9]^. The benefit-cost ratios of such interventions have been estimated as approximately 18:1 for stunting reduction, 4:1 for preschool education, and 3:1 for home visits for underdeveloped children ^[8]^. The potential societal benefits significantly outweigh the costs, making such interventions highly advantageous, particularly in underdeveloped settings. Therefore, it is both imperative and highly effective to intervene during this early childhood period to reduce exposure to risks, with the aim of improving health and cognitive development outcomes in the short term, as well as averting the adverse impacts on health, education, and economic outcomes later in life ^[3,10]^.

While most at-risk children are concentrated in low- and middle-income countries (LMICs), these hazards may be generated or exacerbated by socio-cultural practices and economic constraints, irrespective of geographic location ^[4]^. South Asia and Sub-Saharan Africa have the highest proportion of children under five at risk, at approximately 53% and 66% respectively ^[1]^. These statistics, along with similar estimates published in 2007, have directed many early childhood interventions to these contexts ^[4, 11]^. However, minority groups facing similar socio-cultural norms and economic disadvantages in high-income countries have been largely overlooked to date ^[12]^.

Evidence suggests that a significant proportion of the minority ethnic population in the UK, especially those of South Asian heritage, experience substantial social and economic disadvantage, which may contribute to poorer health outcomes ^[13,14]^. Children from South Asian families residing in the UK are susceptible to many of the same developmental risks as young children residing in South Asia, including poorer birth outcomes and nutritional status ^[15,16]^. The causes of these risks in the UK context are complex but include socioeconomic deprivation, discrimination, language difficulties, cultural norms, and a lack of access to health information, among other factors ^[17]^. Early childhood interventions tailored specifically to these minority ethnic groups can improve health outcomes in early childhood and may avert lifelong disparities in health, education, and economic outcomes. Unfortunately, the availability of such interventions is limited, and there is a need for additional testing of targeted and effective interventions that can be delivered at low cost ^[18,19]^.

The Nurture Early for Optimal Nutrition (NEON) programme was designed to promote equitable early childhood development by educating mothers of South Asian origin in East London on optimal feeding, care, and dental hygiene practices ^[12]^. The intervention is delivered via participatory learning and action (PLA) cycles with women’s groups, which involves active participation, learning, and action by community members and mothers to identify and address problems together. Similar interventions in LMICs, including Bangladesh, Pakistan, and India, have been shown to be effective and cost-effective in reducing maternal and neonatal mortality, improving infant feeding, hygiene, care practices, and thereby enhancing children’s cognitive, language, and motor development outcomes ^[20–24]^. However, there is limited evidence of their success in disadvantaged areas of high-income countries. By adapting this approach for implementation in South Asian communities in East London, the NEON trial will add evidence to the effectiveness and feasibility of PLA in high-income settings. This study conducts a cost analysis of the NEON programme and evaluates its financial sustainability.

## Methodology

### Intervention Design

The NEON Pilot Feasibility Randomised Controlled Trial was conducted from December 2019 to May 2023 to evaluate the feasibility of a definitive trial ^[12, 25]^. A total of 12 clusters, specifically borough wards, were equally randomised to an online treated arm, a face-to-face treated arm, and a control arm. This design balanced the risk of participant contamination while ensuring optimal representation of the South Asian population in East London, including individuals of Indian, Pakistani, Sri Lankan, and Bangladeshi descent.

To enhance the accessibility and feasibility of the programme, Community Facilitators (CFs) from the targeted ethnic backgrounds, Health Visitors (HVs), General Practitioners (GPs), and midwives at each study ward were recruited and involved in the development and delivery of the intervention ^[26]^. The recruitment of participants commenced in May 2022, and implementation began in September 2022. A total of 263 mothers of infants under 24 months from two East London boroughs, Tower Hamlets (TH) and Newham (NH), were enrolled.

Participants in the treated arms received one Participatory Learning and Action (PLA) session every two weeks for a duration of 14 weeks and were followed up for six months afterwards. They were provided with an intervention toolkit, which included picture cards detailing recommended feeding, care, and dental hygiene practices, healthy infant cultural recipes, participatory Community Asset Maps, and a list of resources and services supporting infant feeding, care, and dental hygiene practices ^[25]^. The control group received the usual care under the Healthy Child Programme 0-5 commissioned to local authorities, including regular mandatory postnatal visits and optional prenatal visits ^[12, 27]^. Single blinding was implemented for participant recruitment and outcome assessment. The design and implementation details of the NEON programme will be published separately.

### Costing Method

An economic costing was conducted from the provider perspective. Data were retrieved from the expenditure and accounting records in a trial-specific data collection Excel-based tool ^[12]^. A stepdown procedure was followed to first identify all the costs incurred from the initiation of the trial development to the completion of evaluation and dissemination ^[2]^. Costs associated with start-up, implementation, and monitoring and evaluation activities were distinguished. However, the monitoring and evaluation costs are too intricately entwined to be separated from the implementation costs. They are included as implementation costs as they are likely to be essential for successful programme delivery ^[28]^.

Costs were categorised as capital costs and recurrent costs (staff costs, materials, and consumables). Capital costs include all goods and services having a useful life of more than one year, mainly including computers and cameras. They were depreciated at a rate of 20% per year to account for the value consumed during the trial period ^[29]^. Staff costs mainly include salaries for the study team who were responsible for designing and implementing the NEON intervention, which included activities such as coordinating local HVs, GPs, and Midwifery teams, and recruiting participants and CFs. Other goods and services are deemed as materials such as PLA group facilitator manuals, intervention toolkits, rented venues and snacks for PLA meetings, staff training courses, vouchers for CFs, and overheads. These were valued at the original purchasing value. All costs were inflated or deflated to 2022 values. Results are reported both in 2022-pound sterling (£) and in 2022 international dollars ($INT).

An affordability analysis was conducted by comparing the total cost of the trial delivery to the budget of the local authorities. The efficiency of service delivery was evaluated by assessing two-unit costs, the total cost per beneficiary and the implementation cost per beneficiary. The total cost per beneficiary, i.e., the total cost per mother, was calculated as the total cost divided by the number of participating mothers. The implementation cost per beneficiary (mother) was computed as the total cost per mother excluding fixed start-up costs. It reflects the marginal cost of recruiting and treating one more participant.

Variation in service delivery efficiency was investigated by performing deterministic one-way sensitivity analysis on the total cost per mother. The base case reflects the best approximation of expected unit cost. Changes in assumptions and parameters, such as the number of effectively treated participants, the appropriate capital cost depreciation rate, and the joint cost allocation, were individually assessed against the base case to gauge their influence on service efficiency. Firstly, the number of mothers who proceeded to the first PLA meeting was used as the denominator in the base case. The unit cost variation was explored by using the number of mothers recruited to compute the intention-to-treat (ITT) unit cost. The number of mothers who completed all six PLA meetings was also explored as the denominator, under the stricter assumption of only those women being effectively treated. Second, the capital good depreciation rate was increased and decreased by 10 percentage points to account for the different degree of wear and tear of equipment. Third, variations were introduced to the joint cost allocation, varying the proportion of shared staff, capital, and other recurring costs assigned to PLA implementation and other undertakings like monitoring, evaluation, or research. By adjusting the initial allocation up and down by 20 percentage points, a range of unit costs was derived. This reflects the shifting importance of various activities in future scale-up trials and replications in different contexts. The former will require increased monitoring and evaluation efforts, but the latter should put a greater emphasis on research.

## Results

### Cost Analysis

The trial was conducted in conjunction with the standard care, resulting in all costs being incremental. The total incremental cost of delivering NEON, as depicted in Table 1, across two boroughs in East London amounted to £75,992 ($INT 114,445). The composition of the total cost is as follows: 45% constitutes the salaries for staff responsible for management, supervision, and coordination; 50% is attributed to material costs (Table 1-Panel 1). The capital investment is minimal (5%) and is primarily associated with the electronic equipment required for advocacy, monitoring, and evaluation (Table 1-Panel 1).

**Table 1:** Programme Costs by Component.

| **Table 1: Programme Costs by Component** |  |  |  |
| --- | --- | --- | --- |
| Cost component | Amount (2022 £) | Amount (2022 \$INT) | % of Total cost |
| **Panel 1- Total Cost:** | **75,992** | **114,445** |  |
| *Staff costs* | *34,138* | *51,412* | *45%* |
| *Materials* | *37,979* | *57,198* | *50%* |
| *Capital costs* | *3,875* | *5,836* | *5%* |
| **Panel 2- Cost by Stage:** |  |  |  |
| **Start-up** | **42,949** | **64,683** | **57%** |
| *Staff costs* | *13,238* | *19,937* | *17%* |
| *Materials* | *28,921* | *43,556* | *38%* |
| *Capital costs* | *790* | *1,190* | *1%* |
| **Implementation** | **33,042** | **49,762** | **43%** |
| *Staff costs* | *20,900* | *31,475* | *28%* |
| *Materials* | *9,058* | *13,642* | *12%* |
| *Capital costs* | *3,084* | *4,645* | *4%* |
| **Panel 3- Cost by Site:** |  |  |  |
| Tower Hamlets | 16,470 | 24,804 | 57 women recruited |
| Newham | 59,522 | 89,641 | 206 women recruited |

Out of the total 45% staff costs, 17% were incurred at the start-up stage while 38% were incurred during the implementation stage. Of the total 50% material costs, 38% were expensed at the start-up stage while 12% were expensed during the implementation stage (Table 1-Panel 2).

In total, the start-up cost of £42,949 ($INT 64,683) accounts for 57% of the total cost, while the implementation cost of £33,042 ($INT 49,762) accounts for the remaining 43% (Table 1-Panel 2).

Of the 263 women enrolled, only 186 proceeded to the first PLA meeting and benefited from the intervention of nurturing care. The cost per beneficiary was estimated after accounting for these dropouts. The average cost per mother was estimated to be £409 ($INT 615) (Table 2). Excluding the fixed start-up costs, the implementation cost per mother dropped to £178 ($INT 268), indicating that an additional $INT 268 is required to recruit one more respondent, involve her in the PLA women’s groups, and monitor her behaviour change and children’s health status (Table 2).

**Table 2.** Cost-Efficiency Indicators.

| **Table 2. Cost-Efficiency Indicators** |  |  |
| --- | --- | --- |
| | **Amount (2022 £)** | **Amount (2022 \$INT)** |
| Total cost per mother/beneficiary | 409 | 615 |
| Implementation cost per mother/beneficiary | 178 | 268 |

Pro rata to the number of women recruited in each borough, the total cost of delivering this trial in Newham was estimated at £59,522 ($INT 89,641) (Table 1-Panel 3). This accounts for approximately 0.053% of the borough’s children’s development expenditure in the 2020-21 fiscal year ^[30]^. The proportion was estimated at 4% to cover all 10,967 children under the age of 2 in Newham, if every child has a caregiver undergoing the intervention. In Tower Hamlets, the total trial cost was £16,470 ($INT 24,804), equivalent to approximately 0.003% of Tower Hamlets’ spending on children’s development during the same period ^[31]^.

### Sensitivity Analysis

The sensitivity analyses of the total cost per mother are reported in Table 3. The total cost per mother shows notable sensitivity (changes ranging from −29% to +22%) to the number of participants, i.e., coverage. Using the number of women who proceeded to the first PLA meeting (186 mothers), we calculated the base case total cost per mother at $INT 615. Since start-up costs (57%), as well as the staff and capital costs during the implementation stage (40%) (Table 1-Panel 2), had been budgeted for the total of 263 mothers enrolled, we computed the intention-to-treat (IIT) total cost per mother on wider coverage (263 mothers) at $INT 435 (Table 3). On the other hand, under the more stringent assumption of treatment completion, that only 153 mothers who completed all PLA meetings qualify, the average cost per mother escalates to $INT 748 and rises by 22% (Table 3) relative to the base case.

**Table 3:**
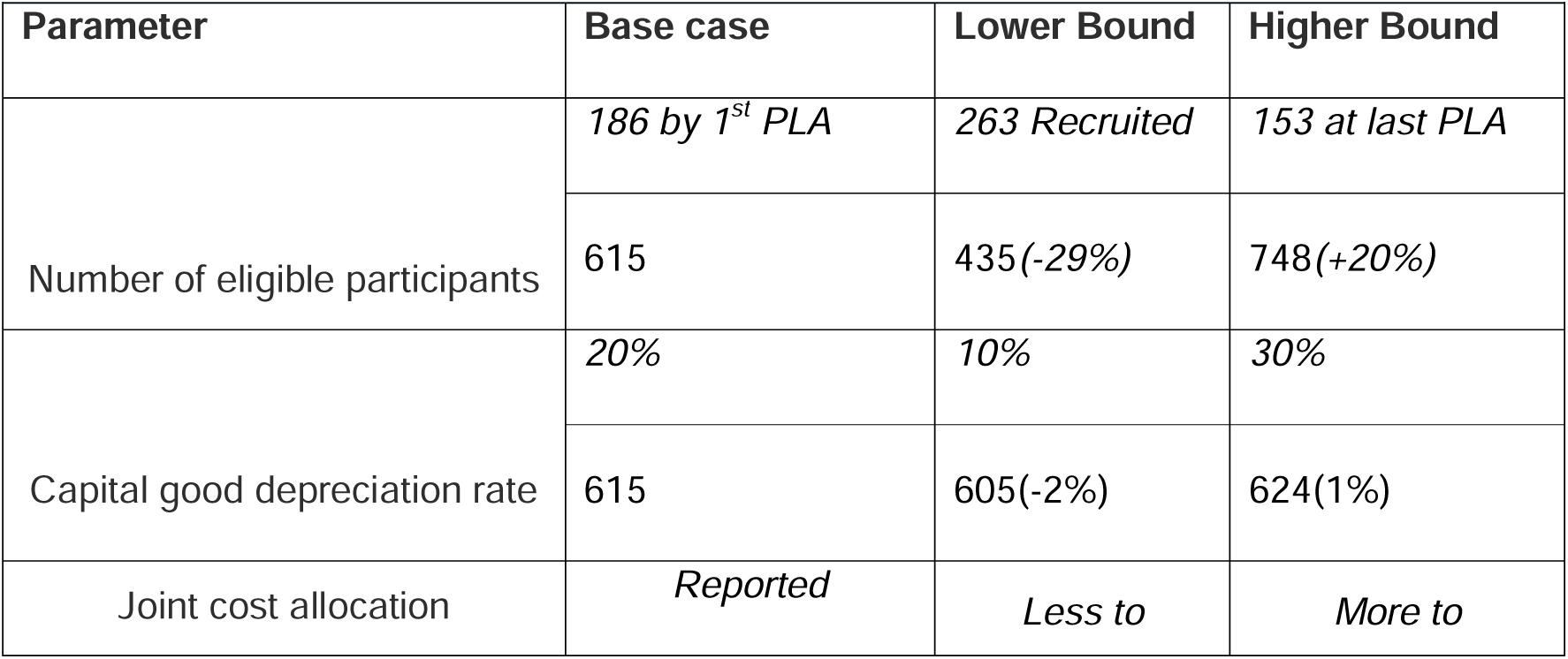
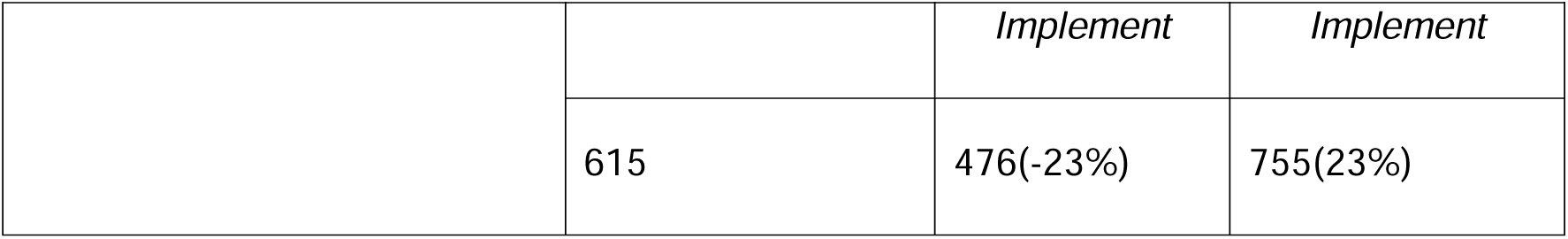
Sensitivity Analysis on total cost per mother($INT)

| Table 3: Sensitivity Analysis on total cost per mother(\$INT) | | | |
| --- | --- | --- | --- |
| Parameter | Base case | Lower Bound | Higher Bound |
| Number of eligible participants | 186 by 1<sup>st</sup> PLA | 263 Recruited | 153 at last PLA |
|  | 615 | 435(-29%) | 748(+20%) |
| Capital good depreciation rate | 20% | 10% | 30% |
|  | 615 | 605(-2%) | 624(1%) |
| Joint cost allocation | Reported | Less to | More to |
|  |  | *Implement* | *Implement* |
|  | 615 | 476(-23%) | 755(23%) |

The total cost per mother is insensitive to the capital good depreciation rate (changes ranging from −2% to 1%), as capital expense only accounts for a small portion of the total cost. As the depreciation rate increases from 10% to 30%, the average cost increases from $INT 605 to $INT 624 (Table 3).

The analysis also reveals that the unit cost is moderately sensitive to the joint cost allocation. Increasing or decreasing the allocation to implementation activities by 20 percentage points would raise or reduce the total cost per mother by 23%.

## Discussion

The objective of this study is to evaluate the costs and affordability of the NEON feasibility trial, an early intervention and life-course approach delivered via Participatory Learning and Action (PLA) cycles with women’s groups, aimed at enhancing the health of South Asian infants in London. The total cost of the trial is £75,992 ($INT 114,445), which is less than 0.1% of the annual child development expenditure of local authorities. If all children under the age of two in one of the boroughs were to be covered, the total cost would be approximately 4% of the borough’s annual child development expenditure.

Given the limited number of studies that have incorporated cost analysis and assessed the cost-effectiveness of PLA women’s groups in improving early childhood nutrition and development globally, this costing study provides additional evidence in this field ^[23,32,33]^. The findings of this study will inform future early childhood interventions via community-led PLA in both low- and middle-income countries (LMICs) and high-income countries.

The incremental start-up cost (INT 64,683), which accounts for more than half of the total cost, is anticipated to be lower for future replicated or scaled-up trials. Necessary groundwork, such as evidence-based implementation strategies, staff training, and community partnership establishment, has already been explored and streamlined ^[17]^. Insights, methodologies, and resources from previous projects can facilitate smoother project initiation, reduce trial-and-error phases, and optimise resource allocation, thereby reducing start-up efforts. Additionally, as the coverage of the intervention increases, economies of scale may come into play. Economies of scale occur when the unit cost decreases as the service delivery increases. In the context of community-based women’s groups, higher coverage indicates that costs such as training, coordination and group facilitation costs, and overheads can be further dispersed among increased participants, resulting in a lower unit cost. A similar intervention conducted in rural India on a larger scale (1253 women covered) estimated their average cost per mother to be INT302 ^[23]^. Though the main drivers of the cost gap are differences in price and cost of living, economies of scale also play a significant part.

However, economies of scale may diminish in potency as coverage is extended to most of the population. The attempt to reach traditionally underserved and marginalised groups may require additional effort ^[10]^. These groups typically face greater barriers to accessing the programme due to their socioeconomic status, geographical location, disability, or other factors. Ensuring coverage to them may require additional actions, resources, and strategies beyond what might be necessary for more privileged or accessible groups. Research on scaling up nutrition interventions has found that when expanded to encompass 80% of the population, the unit cost remained consistent. However, challenges arose when attempting to reach the population between the 80% and 90% coverage range. In this narrower bracket, covering those 10% necessitated a unit cost three to four times higher than the earlier cost (ibid).

Integrating new interventions within existing services can ease some challenges of implementation at scale and therefore be cost saving, but it does carry some risks in maintaining service coverage and quality ^[8,20,21]^. The NEON intervention, implemented via PLA women’s groups, is founded on a community-led model that integrates into the regular care delivery systems of local communities. This approach allows the incorporation of the local context such as the culture, beliefs, and existing practices, potentially leading to boosted accessibility and acceptability ^[8,12]^. By leveraging existing personnel and public resources, the intervention obviates the need for recruiting specialised staff and procuring relevant resources and can be efficiently replicated on a larger scale. Nevertheless, the extensity and quality of service delivery could be largely dependent on the capabilities of the local health system ^[8]^. Limited resources and weak public health systems in other contexts may result in marginalised populations remaining underserved and undermine the central goal of promoting equitable early childhood development. Integrating new interventions into the health system might place additional workloads on health workers, potentially adversely affecting their well-being, performance, and the overall quality of care they provide ^[34]^.

Costs are also sensitive to contexts. Staff salaries, accounting for a considerable share of the total cost (45%), can be reduced if the trial is expanded to or replicated in other regions with a lower average salary than London. Additionally, nearly half of the total cost has been spent on monitoring and evaluation. For feasibility evaluation in this trial, special care has been taken to ensure that interventions are implemented correctly, and frequent and detailed assessments have been carried out to identify potential issues at each stage. These efforts can be reduced in the future if we establish the validity of the invention and construct a proven system to implement it.

The total cost was also somewhat inflated due to the impact of the COVID-19 pandemic. The overall trial duration has been extended to accommodate interruptions caused by lockdowns and sick leaves ^[35]^. Stringent infection prevention and control measures have been implemented, such as enhanced cleaning and disinfection protocols, increased use of Personal Protective Equipment (PPE), and modifications to facility layouts. These situations increased the operational costs. However, the increase is expected to be insignificant since remote working and online PLA meetings were carried out.

## Conclusion

The delivery of NEON is largely within local authorities’ budget for childhood development. The unit cost is sensitive to coverage and joint cost allocation across activities. It is expected to decrease further when sharing costs are spread across more participants and monitoring and evaluation efforts are reduced with validated and well-developed implementing systems in place in future scaled-up trials.

## Data Availability

All data produced in the present study are available upon reasonable request to the authors

## Acknowledgements

The authors would like to thank the South Asian community facilitators in the NEON Intervention and community members of the London Boroughs of Tower Hamlet and Newham for their important contribution and engagement to this research project. Furthermore, the authors would like to acknowledge the contribution of the NEON core team, steering team and all health experts in validating intervention NEON toolkit. We would like to thank the Women & Children First Charity and First Steps Nutrition Trust for their valuable contributions and guidance throughout the study. Members of the NEON Steering Team consist of Prof Atul Singhal, Prof Mitch Blair, Dr Sonia Ahmed, Amelie Gonguet, Gary Wooten, Dr Ian Warwick, Vaikuntanath Kakarla, Prof Richard Watt, Prof Audrey Prost, Dr Edward Fottrell, Ashlee Teakle, Prof Oyinlola Oyebode, Keri McCrickerd, Dr Rana Conway, Professor Lisa Dikomitis, Mari Toomse-Smith, Scott Elliot, Julia Thomas, Aeilish Geldenhuys, Chris Gedge, Kristin Bash, Dr Dianna Smith, Kate Questa, Dr Megan Blake, Gary Tse, Dr Queenie LAW Pui Sze, Gavin Talbot, Dr Chiong Yee Keow Angela Trude Lindsay Forbes Nazanin Zand, Lakmini Shah, Lily Islam, Geromini, Jasvir Bachu, Sumire Fujita, Dina Mobashir, Natasha Chug, Tala Khatib and Delaney Douglas-Hiley.

The NEON Steering Team members had an opportunity to critically review results and contribute to the process of finalising this paper.

The authors would like to thank the National Institute of Health Research, Collaboration for Leadership in Applied Health Research and Care North Thames for funding the NEON study. This work is supported by the NIHR GOSH BRC. The views expressed are those of the author(s) and not necessarily those of the NHS, the NIHR or the Department of Health.

## Author’s Contributions

YZ and PP contributed to the manuscript writing, YZ, PP and SC prepared it for submission. YZ, PP, NB and LM had primary responsibility for the final content. All authors read and contributed to reviewing the study data, the designing of the manuscript, and the approval of the final manuscript.

## Disclaimer

The views expressed in the publication are those of the author(s) and not necessarily those of the sponsor (UCL), funder (NIHR), study partners (Tower Hamlets GP Care Group, London Borough of Newham Council).

## Participant consent for publication

Participant information sheets and consent forms were provided to community members and those expressing interest. Community participants agreed to participate and gave audio/video consent prior to their participation in the workshops. Verbal consent was witnessed and formally recorded. All participants were informed of their right to freely withdraw from the study at any time. Confidentiality of personal data was ensured through the use of anonymisation techniques as stated in the Data Protection Act (1998) and in line with the General Data Protection Regulation (2018). All participant data is anonymised and stored on an encrypted password protected computer. Data can only be accessed by the authorised research personnel.

## Ethics approval

This study has obtained ethical approval from UCL Research Ethics Committee [Ethics ID 17269/001], Sponsor reference number: 142600, Funding Reference: NIHR300020 and IRAS number: 296259, Ref: 21/SW/0142. Study protocols and relevant documents were reviewed by UCL Research Ethics Committee, NHS Health Research Authority (HRA) and study partners involved in establishing data sharing agreement for linking participants’ routine data (Tower Hamlets GP Care Group, London Borough of Newham Council).

## Provenance and Peer Review

Not commissioned; peer reviewed for ethical and funding approval prior to submission.

## Data Sharing Statement

The data supporting the findings of this study is available upon reasonable request from the corresponding author.

## Key Points

- This paper performs a cost analysis to understand the cost implications of running a participatory learning and actions intervention in London.
- This approach allows the incorporation of the local context such as the culture, beliefs, and existing practices, potentially leading to boosted accessibility and acceptability.
- This study deems that the NEON programme on evaluation is a financially sustainable model within the target population.

